# Nerve excitability measured with the TROND protocol in amyotrophic lateral sclerosis: a systematic review and meta-analysis

**DOI:** 10.1101/2022.02.11.22270866

**Authors:** Anna Lugg, Mason Schindle, Allison Sivak, Hatice Tankisi, Kelvin E. Jones

**Affiliations:** Faculty of Kinesiology, Sport, and Recreation, University of Alberta, Edmonton, Alberta, Canada; University of Alberta Library, Edmonton, Alberta, Canada; Department of Clinical Neurophysiology, Aarhus University Hospital, Aarhus, Denmark; Neuroscience and Mental Health Institute, Edmonton, Alberta, Canada

**Author notes:** Correspondence: Kelvin E. Jones, PhD.

**Keywords:** axonal excitability, diagnostic test, electrophysiology, human, ion channels, motor neuron, nerve conduction

## Abstract

**Objective:** This meta-analysis assessed the 30+ nerve excitability indices generated by the TROND protocol to identify potential biomarkers for ALS.

**Methods:** A comprehensive search was conducted in multiple databases, to identify human studies that tested median motor axons. Forest Plot analyses were performed using a random-effects model to determine the pooled effect (Z-score), heterogeneity (I^2^), and Cohen’s d for potential biomarker identification.

**Results:** Out of 2866 studies, 23 studies met the inclusion criteria, incorporating data from 719 controls and 942 ALS patients. Seven indices emerged as potential biomarkers: TEd 90-100 ms, strength-duration time constant (SDTC), superexcitability, TEd 40-60 ms, resting I/V slope, 50% depolarizing I/V, and subexcitability (ranked by the magnitude of the difference between patients and controls from largest to smallest). In a sensitivity analysis focusing on patients with larger compound muscle action potentials (CMAP), only four indices were potential biomarkers: TEd 10-20 ms, TEd 90-100 ms, superexcitability, and SDTC.

**Conclusion:** Among the extensive range of 30+ excitability indices generated by the TROND protocol, we have identified seven indices that effectively differentiate ALS patients from healthy controls. Furthermore, a smaller subset of four indices shows promise as potential biomarkers when the CMAP remains relatively large. However, most studies were considered to be at moderate risk of bias due to case-control designs and absence of sensitivity and specificity calculations, underscoring the need for more prospective diagnostic test-accuracy studies with appropriate disease controls.

**NEW & NOTEWORTHY:** This meta-analysis uncovers seven potential axonal excitability biomarkers for lower motor neuron pathology in ALS, shedding light on ion channel dysfunction. The identified dysfunction aligns with the primary pathology – protein homeostasis disruption. These biomarkers could fill a gap to detect pre-symptomatic spread of the disease in the spinal cord and monitor treatments targeting protein homeostasis and limiting spread, towards enhancing patient care.

## INTRODUCTION

Amyotrophic lateral sclerosis (ALS) is a fatal neurodegenerative disorder characterized by the progressive degeneration of both upper motor neurons (UMNs) and lower motor neurons (LMNs) (1). The clinical manifestations of ALS exhibit significant variability, depending on the degree of UMN and LMN involvement, disease onset location, spread, and the presence of extra-motor symptoms (1–3). The heterogeneity of ALS, coupled with the lack of established biomarkers, poses challenges in diagnosis, prognosis, disease surveillance and therapeutic development.

Several candidate biomarkers have been identified, including: genetic testing, biological fluid analysis, neuroimaging and neurophysiology studies (4–6). Genetic testing provides valuable insights for personalized genetic therapies but has limited utility in monitoring ongoing treatment response (2, 7). Candidate biomarkers for UMN pathology include: neurofilaments, magnetic resonance imaging (MRI), and transcranial magnetic stimulation (TMS). Elevated neurofilaments in cerebrospinal fluid and plasma have susceptibility, prognostic and pharmacodynamic monitoring potential in ALS, but they are not helpful in monitoring focal anatomical spread (8). MRI studies quantify neurodegeneration in ALS, with certain regions showing a correlation with shorter survival (9–11). TMS studies have identified cortical dysfunction as an important early and specific biomarker in ALS (12, 13). For LMN pathology, candidate biomarkers include fasciculations quantified using high-density surface electromyography, ultrasound, and nerve excitability (14, 15). The recently published consensus guidelines for nerve excitability, using the TROND protocol, reaffirm its utility in quantifying LMN pathology (16).

Nerve excitability, assessed using the TROND protocol, provides valuable insights into the biophysical properties of axonal membranes and ion channel function, yielding approximately 30 different excitability indices distributed across five subtests: stimulus-response (SR) curve, strength-duration properties, recovery cycle (RC), threshold electrotonus (TE), and the current-voltage (I/V) threshold relationship (16). Notably, ALS was the first pathophysiology for which this particular nerve excitability test was employed, consistently revealing pathological changes in sodium (Na^+^) and potassium (K^+^) channels (17–23). However, a recent study suggests that approximately 99% of the differences between axons affected by ALS and healthy controls could be attributed to a non-selective reduction in the expression of all ion channels, not exclusively Na^+^ and K^+^ channels (24).

With approximately 30 measures generated from a single TROND nerve excitability test, the crucial first step is to determine which excitability indices effectively differentiate ALS patients from healthy controls and hold potential as LMN biomarkers.

Currently, only narrative reviews and qualitative syntheses comparing the TROND nerve excitability assessment in ALS and controls are available. Therefore, we conducted a systematic review and meta-analysis of existing research to determine: which excitability indices distinguish ALS patients from healthy controls, the reliability and magnitude of those differences, and whether the same indices are appropriate when individuals with ALS have a relatively large compound muscle action potential (CMAP).

## MATERIALS AND METHODS

A systematic review and meta-analysis were conducted following the guidelines of the Preferred Reporting Items for a Systematic Review and Meta-analysis (PRISMA) for Diagnostic Test Accuracy Studies (25) and the Cochrane Handbook for Diagnostic Test Accuracy (26). The search encompassed studies available up to March 12, 2020. Due to circumstances related to the COVID-19 pandemic, the study was not registered with PROSPERO. At the time of registration, PROSPERO indicated significant delays in processing non-COVID-19-related protocols, leading us to proceed with data extraction.

Protocols that have started data extraction are ineligible for PROSPERO registration. Nevertheless, we have taken extra steps to ensure the robustness and transparency of our review process. To support this, we have created an extensive public data repository accessible at https://doi.org/10.6084/m9.figshare.14371904.

### Inclusion and Exclusion Criteria

#### Types of Studies

We considered studies that compared motor axon excitability in the median nerve (abductor pollicis brevis (APB) muscle) using the TROND protocol in people diagnosed with ALS and healthy controls. Reporting of all excitability measures generated by the TROND protocol was not obligatory for inclusion in the meta-analysis. We excluded narrative reviews, theoretical or modelling studies, and animal studies. Additionally, studies not published in English were excluded, while peer-reviewed articles and published dissertations were eligible for inclusion. Abstracts were excluded from the analysis.

#### Participants

The study participants included individuals diagnosed with ALS according to the El Escorial (27), revised El Escorial (28), or the Awaji Criteria (29) along with healthy controls. Both familial ALS (fALS) and sporadic ALS (sALS) cases were included. All ALS phenotypes were included (i.e., bulbar-onset versus limb-onset). Quality assessment included evaluation of the screening of healthy controls participants.

#### Index Test

The index test was the nerve excitability test, administered according to the standardized TROND protocol (16). Studies using a TROND protocol developed prior to the 1999 protocol were evaluated for inclusion on a case-by-case basis (30). For readers unfamiliar with the TROND nerve excitability protocol, additional explanations of terms are available in the consensus guidelines (16). As supplementary information, we have provided a glossary of the excitability indices generated by the TROND protocol, accessible at “Definition of TROND excitability indices.pdf” from https://doi.org/10.6084/m9.figshare.14371904.

#### Reference Standard

The reference standard used in this study was the El Escorial (27), revised El Escorial (28), or the Awaji Criteria (29).

### Search Methods for Identification of Studies

A search was executed by an expert searcher/health librarian (author AS) on the following databases: OVID MEDLINE, PubMed Central, EBSCO CINAHL, OVID EMBASE, OVID HealthSTAR, Scopus, Web of Science – All Databases, and EBSCO SPORTDiscus. See Appendix A for the strategy used to search MEDLINE. The searches were completed using controlled vocabulary (e.g., MeSH) and key words representing variations of the concepts “amyotrophic lateral sclerosis” and “nerve/axonal excitability.” All searches were performed on March 12, 2020. Detailed search strategies for each database are available in the online supplemental information (i.e. Search Strategy.docx).

### Data Collection and Analysis

#### Selection of Studies

Studies were screened using Covidence systematic review software (Veritas Health Innovation, Melbourne, Australia. Available at www.covidence.org). Search results were exported from the databases directly to Covidence by author AS, with duplicate studies removed during the import.

Any additional duplicates found were removed by reviewers during screening. Two independent reviewers (AL and MS) screened titles and abstracts of all identified studies, with disagreements resolved by a third reviewer (KJ). Included citations moved on to full-text review, with the full-text articles uploaded to Covidence. Two reviewers (AL and KJ) independently assessed the articles, and conflicts were resolved by a third reviewer (HT) to finalize the article selection. The reviewers were not blinded to study authors, which carries its own risk of bias, but also allowed for identifying published articles using the same data (e.g. healthy control data replicated in different papers).

#### Data Extraction and Management

A data extraction form was developed within Covidence. Two reviewers (AL and KJ) independently extracted the following variables from the included articles: author information, publication year, country of primary correspondence, study design, reference standard, eligibility criteria, number of participants, mean (or median) age of participants, sex ratio of participants, mean (or median) disease duration, medications, and stratification of patients. Data were also extracted for all reported nerve excitability indices. One reviewer (AL) checked the data entered in the form for accuracy and resolved any discrepancies.

#### Assessment of Methodological Quality

Methodological quality was assessed using the Quality Assessment of Diagnostic Accuracy Studies (QUADAS-2) tool (31) within Covidence. Two independent reviewers (HT and KJ) assessed the methodological quality of included studies. The QUADAS-2 tool involves a structured assessment using signaling questions in four domains: (1) patient selection, (2) index test, (3) reference standard, and (4) flow and timing of participants through the study. Each domain was evaluated for the risk of bias, with the first three domains additionally assessed for applicability concerns. A domain for evaluating case-control studies based on the Newcastle-Ottawa Scale (NOS) was added (modified tool available in supplemental Table 8) (32).

#### Statistical Analysis and Data Synthesis

Data were exported from Covidence to Review Manager, Version 5.4 (RevMan 5.4) software (available at https://training.cochrane.org/online-learning/core-software-cochrane-reviews/revman) for statistical analysis. Meta-analysis, using forest plots, was restricted to excitability measures that were reported in at least four studies. A sensitivity analysis of excitability measures reported in ALS patients with relatively large compound muscle action potentials (CMAP) was conducted to explore potential biomarker indicators before overt atrophy of the APB muscle. All statistical analyses were performed using the RevMan 5.4 software, applying a random-effects model to account for statistical heterogeneity and obtain the mean of true effects (33). Mean difference and 95% confidence intervals were calculated for each excitability measure, with statistical significance determined using the Z-test for overall effect (p<0.05). Cohen’s d was calculated for each excitability measure to determine the magnitude of standardized differences in the means of ALS and healthy controls, with indices having a Cohen’s d below 0.2 excluded as potential biomarkers. Heterogeneity was evaluated using the I^2^ statistic, with values of 25%, 50%, and 75% corresponding to low, moderate, and high heterogeneity between studies (34, 35). Excitability indices with heterogeneity below 50% were considered acceptable (36). Pooled means and standard deviations for each of the nerve excitability indices were calculated for ALS patients and healthy controls, with sample size and sex distribution (%male) across studies presented as mean ± SD. Pooled means and standard deviations were also calculated for participant age, disease duration, and revised ALS Functional Rating Scale (ALSFRS-R).

## RESULTS

### Study Selection

From the initial search, a total of 4867 articles were retrieved. After duplicate removal, 2866 titles and abstracts were screened. Following screening, 43 articles underwent full-text eligibility assessment, with 26 articles fitting the criteria for the systematic review, of which 23 were included in the subsequent meta-analysis. A visual summary of the study selection process and reasons for exclusion can be found in Figure 1. Prisma flow chart of study selection and reasons for exclusion. The inclusion and exclusion criteria were determined a priori and the reasons for exclusion of full-text articles is enumerated, with studies reporting the wrong outcomes (e.g. not reporting the results of a TROND nerve excitability test) as the most prevalent reason for exclusion.

**Figure 1.**
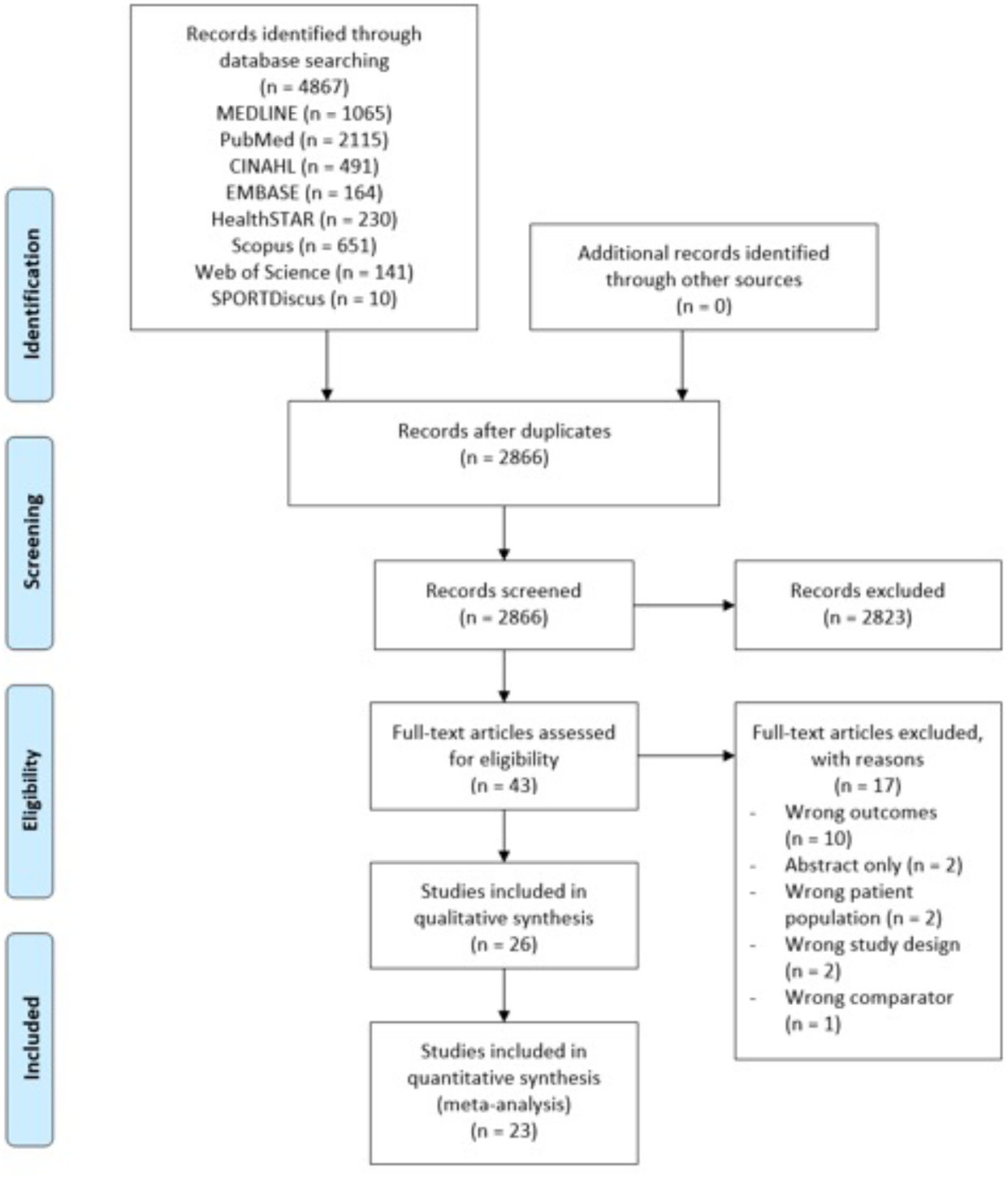
Prisma flow chart of study selection and reasons for exclusion. The inclusion and exclusion criteria were determined a priori and the reasons for exclusion of full-text articles is enumerated, with studies reporting the wrong outcomes (e.g. not reporting the results of a TROND nerve excitability test) as the most prevalent reason for exclusion.

### Study Characteristics

Twenty-six articles consisting of a total of 942 ALS patients and 719 healthy controls were included in the systematic review. Published within the last 25 years (1996 to 2020), these studies were conducted across five countries: Australia (n=14), Japan (n=9), Germany (n=1), Korea (n=1), and Portugal (n=1). All studies used either the El Escorial (n=5), revised El Escorial (n=11), or Awaji (n=5) criteria as the reference standard. Three studies used both the revised El Escorial and Awaji criteria. One study used a combination of genetic testing (*C9orf72*) and Awaji criteria. One study did not report the specific reference standard used. Study characteristics are summarized in supplemental Table 9.

### Participant Characteristics

The included studies exhibited a wide range of sample sizes, as reported in Table 1. ALS patients showed a slightly older pooled mean age compared to healthy controls, with a modestly larger proportion of males in the ALS group (Table 1). Ten studies reported average disease duration (17.7 months, range 12.5 to 25.9) while seven studies reported median disease duration (ranging from 9.5 to 16.1 months). The pooled mean ALS Functional Rating Scale – Revised (ALSFRS-R) score was 40.5 (from 8 studies, range 39.0 to 42.1). A summary of the participant characteristics is provided in Table 1, with further details available in supplemental Table 9.

**Table 1.** Summary of participant characteristics.

|  | ALS |  |  | Healthy Controls |  |  |
| --- | --- | --- | --- | --- | --- | --- |
|  | Mean(SD) | Range | # studies (/26) | Mean(SD) | Range | # studies (/26) |
| **Sample size** | 33.6(30.0) | 6-140 | 26 | 27.6(9.20) | 10-48 | 26 |
| **Mean Age (years)** | 61.4(9.40) | 44.0-68.0 | 17 | 54.6(10.5) | 40.4-66.0 | 13 |
| **Sex (% male)** | 61.6(10.1) | 41-83 | 21 | 51.7(10.8) | 30-72 | 19 |
| **Mean Disease duration (months)** | 17.7(14.9) | 12.5-25.9 | 10 | n/a | n/a | n/a |
| **Mean ALSFRS-R** | 40.5(5.00) | 39.0-42.1 | 8 | n/a | n/a | n/a |

Sample size and sex are represented as mean (standard deviation). Age, disease duration, and ALSFRS-R are represented as pooled mean (pooled SD); the range for these characteristics is the range of the means in the included studies. n/a = not applicable.

### Comparison of Nerve Excitability Indices

Out of approximately 30 TROND nerve excitability indices, sixteen were reported by four or more studies and analyzed. To aid readers unfamiliar with the standard TROND excitability indices and their definitions, we have provided a file with explanations in the online supplemental material (“Definition of TROND excitability indices.pdf” <u>at</u> https://doi.org/10.6084/m9.figshare.14371904).

Among the analyzed indices, ten showed significant pooled effects (Z ranging from 9.88 to 2.81, in descending rank order): depolarizing Threshold Electrotonus at latencies between conditioning and test stimuli of 90-100 ms (TEd 90-100), strength-duration time constant (SDTC), superexcitability, compound muscle action potential (CMAP), depolarizing Threshold Electrotonus at latencies of 40-60 ms (TEd 40-60), depolarizing Threshold Electrotonus at latencies of 10-20 ms (TEd 10-20), resting current/voltage (I/V) slope, threshold change during 50% depolarizing I/V test, subexcitability, and rheobase.

Since CMAP (I^2^ = 88%) and TEd 10-20ms (I^2^ = 51%) exceeded the cut-off of 50% heterogeneity, they were excluded from further consideration. Rheobase fell below the small effect size cut-off based on Cohen’s d (0.2), resulting in its exclusion from further consideration. Consequently, the remaining seven excitability indices are proposed as candidate biomarkers. Table 2 gives a summary of the number of studies reporting each excitability index, heterogeneity, overall effect, p-value, and Cohen’s d. Forest plots for all excitability indices are available in online supplemental figures 1-12, along with pooled means and standard deviations for all extracted nerve excitability indices in supplemental Table 10.

**Table 2.** Summary statistics for axonal excitability indices reported by four or more studies.

| Excitability Index | # Studies reporting index | Heterogeneity ($I^2$) | Overall Effect (Z) | P value | Cohen's d |
| --- | --- | --- | --- | --- | --- |
| TEd 90-100ms | 17 | 38 | 9.88 | <0.00001 | 0.72 |
| SDTC | 21 | 3 | 9 | <0.00001 | 0.44 |
| Superexcitability | 17 | 35 | 8.54 | <0.00001 | 0.60 |
| CMAP Amplitude | 19 | 88 | 8.51 | <0.00001 | 1.2 |
| TEd 40-60ms | 8 | 4 | 5.57 | <0.00001 | 0.52 |
| TEd 10-20ms | 10 | 51 | 4.6 | <0.00001 | 0.28 |
| Resting I/V Slope | 6 | 0 | 4.35 | <0.0001 | 0.44 |
| 50% Depolarizing I/V | 4 | 0 | 4.14 | <0.0001 | 0.42 |
| Subexcitability | 15 | 37 | 4.07 | <0.0001 | 0.29 |
| Rheobase | 11 | 0 | 2.81 | 0.005 | 0.02 |
| TEh 90-100ms | 14 | 24 | 1.93 | 0.05 | 0.13 |
| 100% Hyperpolarizing I/V | 5 | 52 | 1.4 | 0.16 | 0.12 |
| Hyperpolarizing I/V Slope | 11 | 67 | 1.04 | 0.3 | 0.10 |
| RRP | 11 | 0 | 0.59 | 0.56 | 0.01 |
| Refractoriness | 7 | 45 | 0.34 | 0.74 | 0.07 |
| TEh 10-20ms | 4 | 82 | 0.09 | 0.93 | 0.12 |

Indices descending rank ordered (top to bottom) for overall effect (Z). Indices with a P value $\geq 0.5$ have a grey background.

It is worth noting that not all indices generated by the TROND nerve excitability test were reported; there was a bias toward reporting indices that demonstrated significant differences between ALS patients and healthy controls. Measures that were not different between ALS and healthy controls were often reported without raw data and could not be included in the analysis.

#### Compound Muscle Action Potential

Nineteen studies reported maximum CMAP amplitude (ALS = 644, Healthy Control = 533), with marked heterogeneity observed (I^2^=88%) (12, 18, 20, 21, 23, 37–49). As anticipated, CMAP was significantly reduced in ALS patients compared to healthy controls, with a mean difference of −3.58 mV (95%CI: −4.41 to −2.76).

Interestingly, four studies included ALS patients with CMAP values similar to healthy controls, suggesting nerve excitability testing was conducted before overt atrophy of the APB muscle (i.e. no significant differences in CMAP between the ALS and Control groups Z=1.62, p=0.10; I^2^=0%) (12, 19, 21, 46). To explore whether changes in nerve excitability could be detected before a decline in CMAP, we performed a sensitivity analysis using these four studies. The sensitivity analysis revealed that only four of the seven excitability indices proposed as candidate biomarkers in Table 2 showed significant differences when the CMAP values were relatively large. Figures 2 – 5 display the forest plot meta-analysis for these four indices. The forest plot meta-analysis for the remaining twelve TROND nerve excitability indices are available in the supplemental materials.

**Figure 2.**
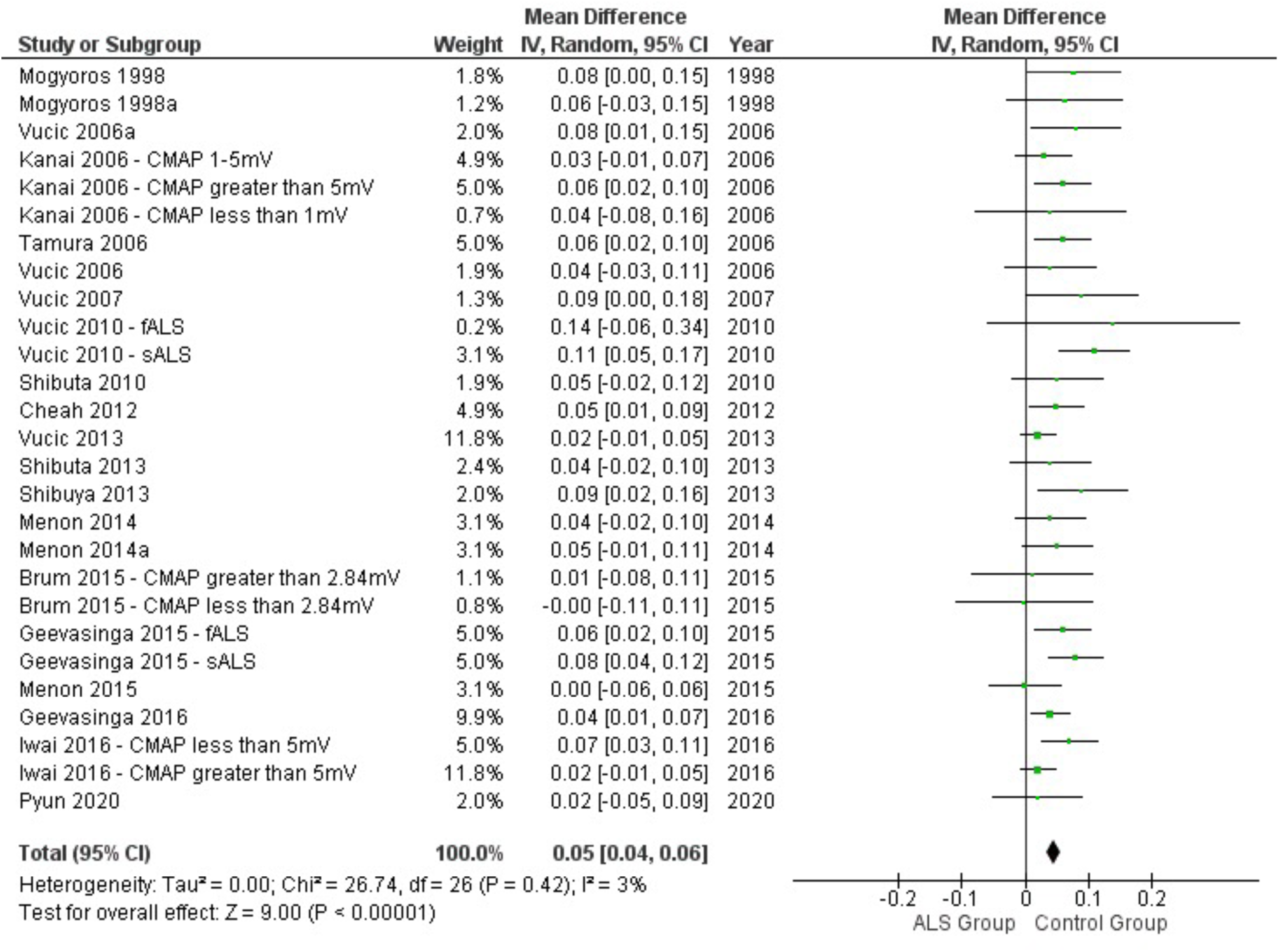
Forest plot of strength-duration time constant (SDTC) in ALS patients and healthy controls. Data from 21 studies, stratified into 27 groups, were analyzed to compare SDTC (ALS = 678, Healthy Control = 558). On an individual group level only 12 of 27 comparisons showed significant differences between the ALS and Control groups. However, the overall mean difference (diamond) was significant with a difference between the two groups of 0.05 ms, or 50 microseconds, meaning SDTC was longer in ALS patients. The sensitivity analysis found SDTC was significantly different even when patients with ALS have a relatively large CMAP compared to healthy controls (Z=2.16, p=0.03). An increase in SDTC is often associated with a stronger contribution of persistent sodium channels to axon threshold, but this is not the only biophysical mechanism that influences SDTC.

#### Strength-Duration Properties

Two excitability indices from the strength-duration component of the TROND nerve excitability test were analyzed: strength-duration time constant (SDTC, also known as chronaxie), and rheobase. SDTC was significantly longer in ALS patients compared to healthy controls. SDTC continued to detect a difference between patients and controls when the analysis was restricted to patients with relatively large CMAP values (Z=2.16, p=0.03). Therefore, SDTC has potential as a biomarker prior to overt atrophy of the APB muscle.

Eleven studies reported rheobase (12, 22, 23, 40, 43, 45–50) with an overall mean difference of −0.29 mA (95%CI: −0.49 to −0.09, supplemental Figure 2), showing that rheobase is reduced in ALS patients compared to healthy controls. While the overall effect was significant, the magnitude of the difference was negligible (d=0.02, Table 2) which excluded this excitability index from further consideration.

#### Recovery Cycle

In the recovery cycle component of the TROND nerve excitability test, we analyzed four excitability indices: superexcitability, subexcitability, relative refractory period (RRP) and refractoriness at 2 ms (Table 2). Of these, only superexcitability consistently distinguished ALS patients from healthy controls, both the full data set and when restricted to patients with relatively large CMAP values.

Superexcitability was more negative in ALS compared to healthy controls, indicating a lower threshold in the period of 5-10 milliseconds following a preceding compound action potential (ALS = 624, Healthy Control = 442, Figure 3). Subexcitability was significantly lower in ALS patients compared to healthy controls (ALS = 586, Healthy Control = 407, supplemental Figure 3). However, when analysis was restricted to the subset of patients with relatively large CMAP values, there was no detectable difference in subexcitability between ALS and healthy controls. Relative refractory period (RRP) showed no significant difference between patients and healthy controls in eleven studies (ALS = 283, Healthy Control = 298, supplemental Figure 4). Refractoriness at 2 ms also showed no significant difference between patients and healthy controls in seven studies (ALS = 358, Healthy Control = 175, supplemental Figure 5).

**Figure 3.**
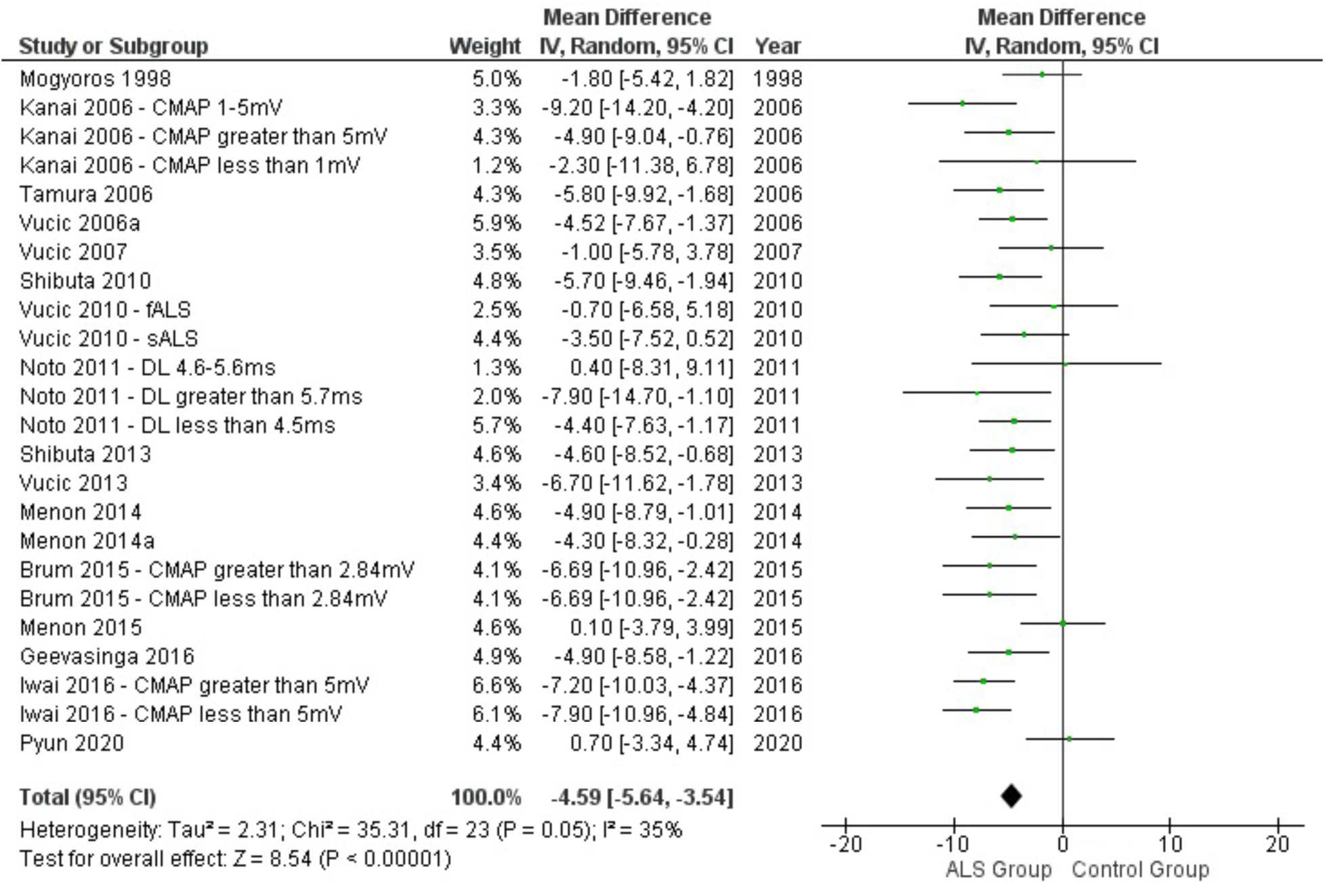
Forest plot of superexcitability in ALS patients and healthy controls. Data from 17 studies were analyzed (ALS = 624, Healthy Control = 442) and a mean difference of −4.59 percent was found. At an individual group level, 16 out of 24 comparisons differentiated between the ALS and Control groups. (Note: superexcitability is the decrease in threshold relative to baseline and is expressed as a percent change.) The sensitivity analysis found that superexcitability was significantly different even when patients with ALS were restricted to the subset with relatively large CMAPs (Z=2.8, p=0.005). This means that at the lowest point of the superexcitable period, the ALS group dipped lower than controls which indicates less current is needed to reach threshold following a preceding action potential. Mechanistically this would be consistent with an increased size of the depolarizing afterpotential in people diagnosed with ALS which may arise from decreased juxtaparanodal fast potassium channel conductance.

#### Threshold Electrotonus

In the threshold electrotonus component of the TROND nerve excitability test, sixteen excitability indices are generated: ten in response to depolarizing conditioning current (TEd) and six in response to hyperpolarizing conditioning current (TEh). However, only five of these indices were sufficiently reported in the literature for analysis. Among these, only two consistently distinguished ALS patients from healthy controls, both in the full dataset and when considering patients with relatively large CMAP values.

Three reported indices generated during depolarizing threshold electrotonus showed significant differences between ALS patients and healthy controls (Table 2). These are ranked by the size of the pooled effect (Z): TEd 90-100ms (Figure 5), TEd 40-60ms (supplementary Figure 6), and TEd 10-20ms (Figure 4). Notably, only TEd 10-20ms and TEd 90-100ms distinguished between healthy controls and ALS patients with relatively large CMAP values.

**Figure 4.**
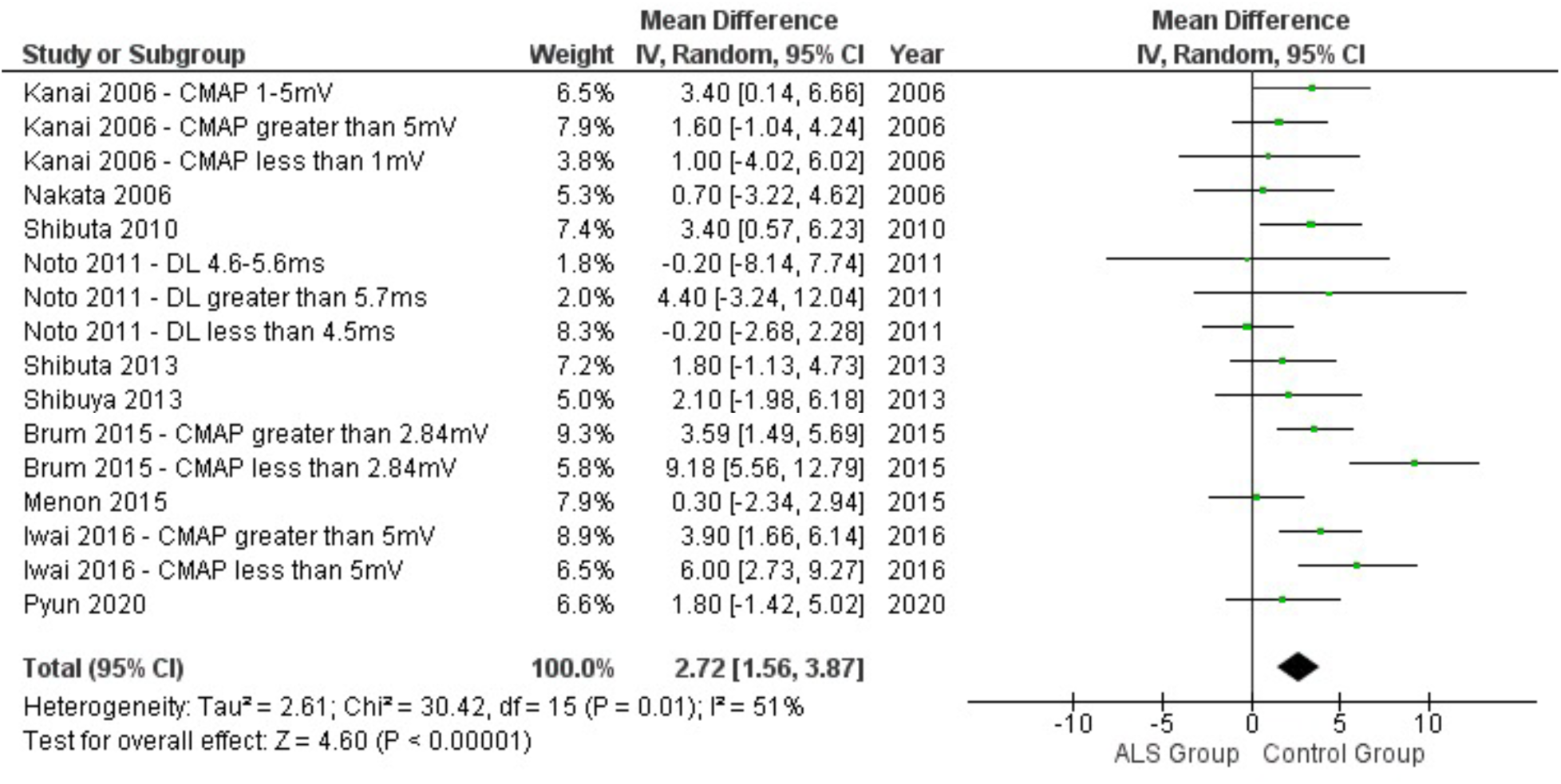
Forest plot of TEd 10-20ms in ALS patients and healthy controls. Data from 10 studies, stratified into 16 groups, were analyzed (ALS = 429, healthy control = 244) and a mean difference of 2.72% (95%CI: 1.56 to 3.87) was found, but with heterogeneity above the threshold of 50% (I^2^=51%). At the individual group level, 6 out of 16 comparisons differentiated between the ALS and Control groups. The sensitivity analysis also found that TEd 10-20ms was significantly different when patients with ALS were restricted to the subset with relatively large CMAPs (Z=2.99, p=0.003) and heterogeneity was moderately improved (I^2^=47%). This means that at early latencies of 10-20 ms following depolarization of the membrane potential from rest, the threshold reduction is about 2.5-2.7% greater in the ALS group. This phase of depolarizing threshold electrotonus is sometimes labelled as the S1 phase, and the fast potassium channels (i.e. K_v_1 family with high density at the juxtaparanode) act to limit the threshold reduction. The higher threshold reduction in ALS patients is consistent with decreased efficacy of the fast potassium channels.

**Figure 5.**
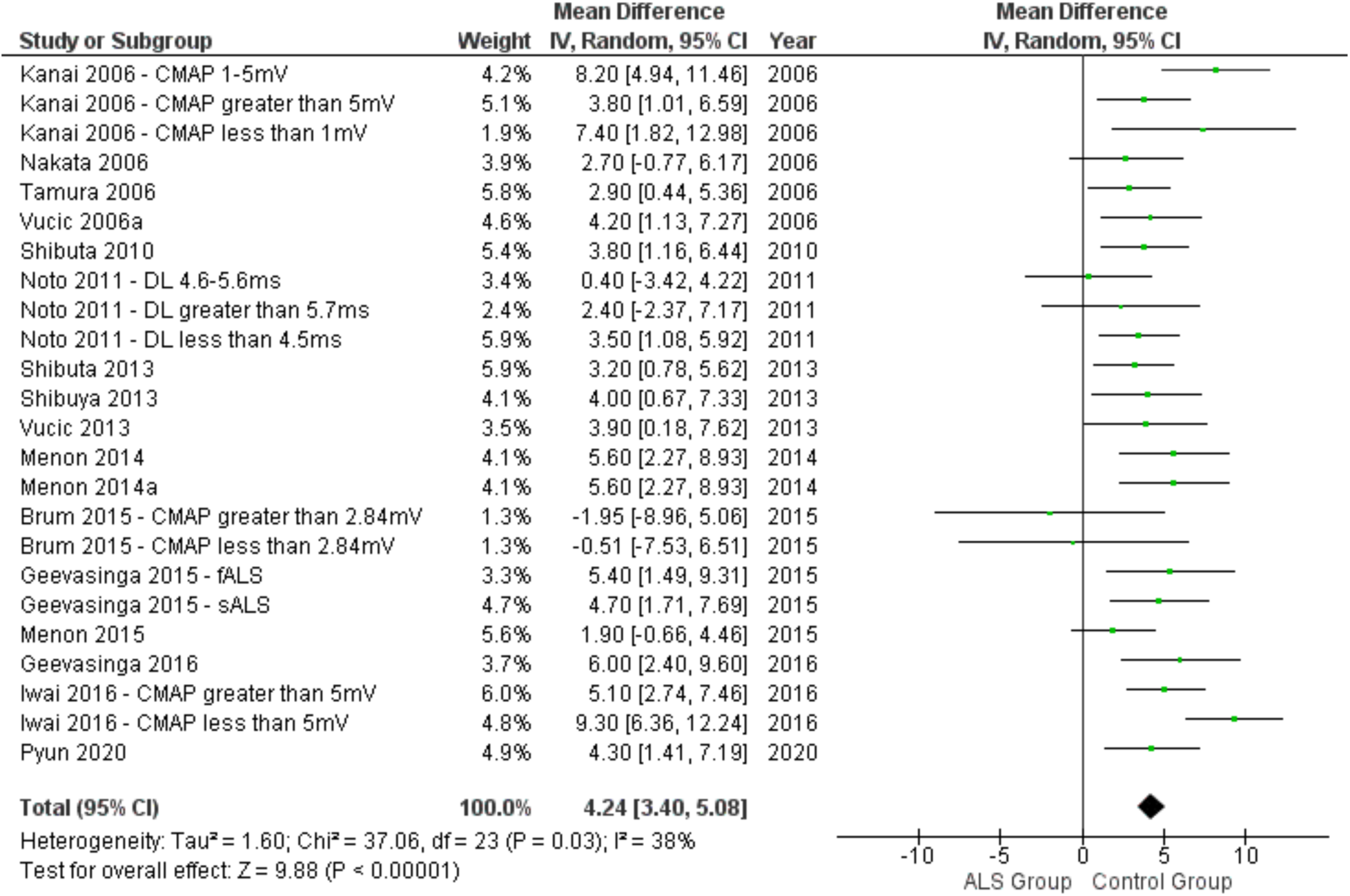
Forest plot of TEd 90-100ms in ALS patients and health controls. Data from 17 studies were analyzed (ALS = 613, healthy controls = 445) and a mean difference of 4.24% (95%CI: 3.40 to 5.08) was found. The Z-value for the overall effect was the greatest for this index (Table 2) and this index was also significant in the subanalysis with ALS patients that have relatively large CMAPs (Z=2.96, p=0.003). At the individual group level, 18 out of 24 comparisons differentiated between the ALS and Control groups. This means that at long latencies of 90-100 ms following depolarization of the membrane potential from rest, the threshold is more excitable in the ALS group compared to healthy controls. This later phase of depolarizing threshold electrotonus is sensitive to both fast and slow potassium channels (K_slow_ channels are from the K_v_7 family) and the threshold difference in ALS patients is consistent with decreased efficacy of both fast and slow potassium channels.

Neither of the two indices generated during hyperpolarizing threshold electrotonus showed significant differences between patients and healthy controls (Table 2): TEh 10-20 ms (supplemental Figure 7), and TEh 90-100ms (supplemental Figure 8).

#### Current-Threshold (I/V)

In this section of the TROND nerve excitability test, we analyzed four indices: resting I/V slope, hyperpolarizing I/V slope, 50% depolarizing I/V, and 100% hyperpolarizing I/V (Table 2). The latter two indices measure the percent reduction of threshold in response to conditioning polarizing currents. Specifically, 50% depolarizing I/V measures the reduction of threshold (%) after a 200 ms subthreshold depolarizing conditioning pulse with an amplitude half that of the unconditioned threshold, indicating outward rectification. Conversely, 100% hyperpolarizing I/V is an index of inward rectification.

From these four indices, resting I/V slope (supplemental Figure 11) and 50% depolarizing I/V (supplemental Figure 9) differentiated ALS patients from healthy controls in the full dataset but not when considering patients with relatively large CMAP values. Neither hyperpolarizing I/V slope (supplemental Figure 12) nor 100% hyperpolarizing I/V (supplemental Figure 10) exhibited any significant differences between patients and healthy controls.

### Methodological Quality

The methodological quality of the included articles (n=26) was assessed using the QUADAS-2 tool, and the itemized results for individual articles are presented in supplemental Table 11. An overview of the author’s judgements across all included articles is provided in supplemental Figure 13. The overall methodological quality of the primary research is moderate. There is a clear need to progress from case-control to prospective, longitudinal studies that include appropriate disease controls rather than healthy controls. Notably, the majority of papers performed group comparison using traditional parametric null-hypothesis significance testing rather than the preferred reporting of sensitivity, and specificity for diagnostic test accuracy studies.

#### Patient Selection

Most studies (n=21) were judged to have a high risk of bias in the patient selection domain, primarily due to non-consecutive sampling and the use of a case-control design. The remaining studies were assessed as unclear (n=3) or low (n=2) risk of bias. To account for the case-control design common in early medical test research, an additional domain was added to the QUADAS-2 tool based on the Newcastle-Ottawa Quality Assessment Scale (NOS). The risk of bias assessment in this domain resulted in: low (n=9), unclear (n=14), or high (n=3) risk of bias judgements. Unclear and high risk assessments were mainly due to inadequate reporting of healthy control selection, screening and exclusion criteria.

#### Index Test

The majority of studies (n=24) were judged to have a low risk of bias for the index test domain, with two studies receiving unclear risk of bias assessments.

#### Reference Standard

In the reference standard domain, most studies (n=23) were judged as having a low risk of bias, while the remaining had unclear (n=1) or high (n=2) risk of bias.

#### Flow and Timing

Most studies (n=16) had low risk of bias in the flow and timing domain, while eight and two studies had unclear and high risk of bias, respectively.

#### Applicability Concerns

Seventeen studies were rated as having low applicability concerns related to patient selection, while eight studies received unclear ratings, primarily due to extended disease durations. A single study was rated as having high applicability concerns due to the use of axonal excitability testing on single axons, leading to its exclusion from the meta-analysis.

## DISCUSSION

The primary objective of this systematic review and meta-analysis was to identify potential lower motor neuron (LMN) electrophysiological biomarkers for ALS from the more than 30 nerve excitability indices generated by the TROND protocol. We identified seven candidate biomarkers using the full data set (719 controls and 942 ALS patients), and four when the ALS participants were restricted to individuals with a relatively large compound muscle action potential (CMAP, 103 controls and 109 ALS patients). The candidate biomarkers come from different subtests of the TROND protocol: Strength-Duration (strength-duration time constant, SDTC), Recovery Cycle (superexcitability and subexcitability), Threshold Electrotonus (TEd 90-100ms, TEd 40-60ms, and TEd 10-20ms from the patients with larger CMAPs), and the Current-Threshold I/V subtest (resting I/V slope, and 50% depolarizing I/V). In this discussion, we aim to assess the clinical implications of these findings, particularly their potential for diagnostic use and disease progression monitoring in ALS. But first we explore how these results contribute to advancing our understanding of the underlying axonal pathology mechanisms in ALS.

### Axon Membrane Pathology

The pathophysiology of axonal ion channels in ALS has been a topic of discussion since the earliest application of the TROND nerve excitability test (17). Previous interpretations pointed to changes in ion channel conductances, particularly Na+ and K+ conductances, in ALS axon pathology (21, 23, 47, 50). However, these inferences, derived from single studies, were not always consistent. Could these inferences withstand quantitative integration through a meta-analytical approach?

The routine measure used to infer voltage-gated Na^+^ channel function in the TROND protocol is the strength-duration time constant (SDTC), which was identified as a candidate biomarker in our meta-analysis (Figure 2). In ALS, the Na_V_1.6 sodium channels, concentrated at the nodes of Ranvier (51, 52), exhibit a persistent inward current alongside the traditional transient current (53–55). The presence of Na_V_1.6 channels operating in the persistent mode alters the relationship between stimulus strength and duration, resulting in a longer SDTC (21–23, 38, 47). The meta-analysis reinforces the ongoing assertion that ALS is associated with changes in axonal Na^+^ channels, indicating an increase in persistent inward current.

Similarly, the TROND protocol assesses multiple excitability indices to infer the function of voltage-gated K^+^ channels. Specifically, slow K^+^ channels, K_V_7 family, concentrated at the nodes of Ranvier (56, 57) and fast K^+^ channels, K_V_1 family, concentrated in the juxtaparanode (58–60) contribute to multiple indices. The meta-analysis identified three potential biomarkers (TEd 90-100ms, TEd 40-60ms, and TEd 10-20ms) consistent with a decrease in both fast and slow K^+^ channels (Figure 5, supplementary Figure 6, Figure 4). The reduced function of K+ channels is further supported by the decrease in outward rectification in ALS patients (measured by the 50% depolarizing I/V value, supplementary Figure 9) and the reduction in subexcitability (supplementary Figure 3). The meta-analysis affirms that ALS is accompanied by decreased function of both fast and slow K^+^ channels.

While individual studies yielded equivocal results, our meta-analysis provided a definitive answer. For instance, out of 27 comparisons of SDTC, only 12 differentiated between ALS and controls, or 44% (Figure 2). However, the meta-analysis demonstrated a significant overall effect, showing low heterogeneity between studies and a medium-sized difference between ALS and controls. These results offer a conclusive answer, resolving uncertainties that persist in narrative reviews of individual studies.

### Peripheral Axons and the Primary Pathology in ALS

The identification of seven candidate biomarkers linked to voltage-gated ion channels shouldn’t be interpreted as evidence that the primary lower motor neuron (LMN) pathology in ALS is a channelopathy. Current understanding suggests that dysfunction in RNA processing, leading to altered protein homeostasis, likely initiates and propagates the pathology (61, 62). Approximately 97% of ALS cases result in cytoplasmic aggregation of the RNA-binding protein TDP-43 (63), leading to atypical RNA splicing and subsequent loss of crucial axon-supporting proteins such as stathmin-2 in the peripheral nervous system (64, 65). Single-nucleus RNA sequencing of human spinal motor neurons has identified a number of genes that characterize these cells related to structure, size and ALS – but voltage-gated ion channels were not a predominate category (66).

In this context, voltage-gated ion channels are merely a category of transmembrane proteins whose homeostasis is disrupted by TDP-43 pathology. However, these proteins are measurable using electrophysiological tools like the TROND protocol. Researchers have already used the TROND protocol together with complementary mathematical modeling to suggest a non-selective reduction in all axonal ion channels that is consistent with a primary TDP-43 pathology (24).

Therefore, the physiological dysfunction observed in axons, as measured by these candidate biomarkers, appears to be a downstream consequence of a primary pathology occurring elsewhere. Consequently, therapeutic interventions aimed at normalizing axon electrophysiology could be compared to treating urinary frequency in individuals diagnosed with diabetes mellitus. The candidate axon biomarkers are indicators of a pathogenic process and could be used to indicate efficacy of therapeutic interventions targeting the upstream pathology.

### Limitations

This study has limitations that warrant acknowledgement. For example, we restricted our analysis to studies published in English, potentially missing some data and introducing a language bias. The existing English language literature predominantly comprised case-control studies comparing axonal dysfunction in diagnosed ALS patients and healthy controls. The absence of disease controls reduces the strength of the conclusions about whether the identified excitability biomarkers are specific to ALS or are indicative of conditions affecting general axonal health. Future diagnostic test accuracy studies should include disease controls and calculate measures of sensitivity and specificity.

Secondly, not all measures generated during a TROND nerve excitability test were reported equally across studies, leading to potential biases in the indices reported. Encouraging the sharing of anonymized individual participant data to facilitate mega-analyses could mitigate this limitation, ensuring a more comprehensive and unbiased analysis of TROND excitability measures in ALS.

Additionally, the presence of repeated patient data in studies by the same authors posed a challenge in the analysis. While we believe the impact of these repetitions on the results was minimal, acknowledging this potential source of bias is essential for a transparent evaluation. This issue could also be mitigated by sharing anonymized individual participant data.

Lastly, our meta-analysis was based on cross-sectional data, preventing an assessment of longitudinal changes in individual patients. Future research should prioritize longitudinal studies to determine the changes of these excitability indices, enabling an understanding of the prognostic implications.

### Clinical Implications

In evaluating the clinical utility of these biomarkers, we adhere to the BEST (Biomarkers, EndpointS, and other Tools) framework established by the U.S. Food and Drug Administration (FDA) and the National Institutes of Health (NIH), following the precedent set by neurofilament light chain (NfL) studies in ALS (8, 67). The multifaceted journey from symptom onset to diagnosis is influenced by diverse factors, resulting in diagnostic delays and initial misdiagnosis for about 40% of patients (68). In the BEST framework, a diagnostic biomarker should ideally expedite diagnosis or pinpoint specific disease subtypes.

Our analysis doesn’t propose that axon excitability biomarkers shorten the diagnostic timeline or circumvent misdiagnosis. Instead, it identifies key indices among the 30+ excitability measures. For patients without over thenar muscle atrophy, the focus narrows to four candidate biomarkers, as revealed in our sensitivity analysis (TEd 10-20ms, TEd 90-100ms, superexcitability, and SDTC). Recent class I evidence supports the diagnostic potential of this subset of four, demonstrating high accuracy in a consecutive case series study involving 216 patients with suspected motor neuron disease (69). The risk score developed in that study, based in part on the preprint of the current manuscript, correctly identified patients with ALS, and more importantly 73% of difficult-to-diagnose cases at baseline, offering the potential for earlier intervention and reduced psychological distress for patients and caregivers (70, 71).

While neurofilament light chain (NfL) proves invaluable for assessing disease aggressiveness and pharmacodynamic responses (8), it falls short in tracking the anatomical spread of LMN pathology from the body region of onset (3). Peripheral motor axon studies indicate that nerve excitability changes precede axonal loss (21, 69), suggesting a potential role for axon excitability biomarkers in monitoring disease progression and guiding interventions. Bilateral TROND nerve excitability testing across limbs emerges as a promising avenue for tracking spread within the spinal cord. However, these speculations necessitate further investigation to determine their clinical utility.

## Conclusions

This study uncovers seven potential ALS biomarkers within the TROND nerve excitability protocol, offering promising avenues for diagnosis, monitoring disease spread, and therapeutic interventions designed to limit spread or restore protein homeostasis in axons. These candidate markers are promising adjuncts to the emerging arsenal of ALS biomarkers.

To fully realize the clinical utility of these markers, future studies must navigate the limitations identified in this meta-analysis. Incorporating disease controls in diagnostic test accuracy studies is paramount, enabling a quantitative understanding of the specificity and sensitivity of these biomarkers in the heterogeneous phenotypes of ALS and its mimics. Encouraging the secure sharing of raw anonymized individual participant data would bridge gaps in reporting, ensuring a comprehensive evaluation of all indices generated during nerve excitability tests.

Moving forward, researchers should concentrate on refining and validating these biomarkers alongside deep clinical phenotyping, genetic analyses, and other emerging biomarkers to explore their potential in longitudinal studies to understand disease progression patterns and therapeutic responses. A focus on standardizing protocols, enhancing collaboration between research groups, and exploring innovative data-sharing platforms could foster a platform from which the future landscape of ALS understanding and treatment can be shaped (72).

## Data Availability

All data used in this meta-analysis are freely available on the Figshare data repository at https://doi.org/10.6084/m9.figshare.14371904. Supplementary material are available at the same DOI.

## APPENDIX

## Appendix A MEDLINE Search Strategy

1. exp Motor Neuron Disease/ or exp Amyotrophic Lateral Sclerosis/
2. (moto$1 neuron$1 disease$1 or moto?neuron$1 disease).mp.
3. ((Lou Gehrig$1 adj5 syndrome$1) or (Lou Gehrig$1 adj5 disease)).mp.
4. charcot disease.tw.
5. Amyotrophic Lateral Sclerosis or als.ti,ab,kf.
6. or/1-5
7. Electric Stimulation / mt [Methods] or chronaxy/ or electromyography / mt
8. (chronaxy or neural conduction).ti,ab,kf.
9. ((axonal excitab*) or (nerve excitab*)).ti,ab,kf.
10. Evoked Potentials, Motor / ph [Physiology] or Axons / ph or Action Potentials / ph or Neural Conduction / ph [Physiology]
11. or/7-10
12. 11 AND 6
13. animals/ not (exp animals/ and humans/)
14. 12 not 13

## DATA AVAILABILITY

All data used in this meta-analysis are freely available on the Figshare data repository at https://doi.org/10.6084/m9.figshare.14371904. The data file is “ExtractedData.rm5”.

## SUPPLEMENTAL MATERIAL

The file “Search Strategy.docx” contains the detailed search data for all citations found from each of the eight different databases (https://doi.org/10.6084/m9.figshare.14371904).

The file “Supplemental Materials.pdf” available from https://doi.org/10.6084/m9.figshare.14371904 contains the following material:

Table 1. Search strategy for PubMed Central

Table 2. Search strategy for CINAHL

Table 3. Search strategy for EMBASE

Table 4. Search strategy for HealthSTAR

Table 5. Search strategy for Scopus

Table 6. Search strategy for Web of Science

Table 7. Search strategy for SportDiscus

Table 8. Modified QUADAS-2 Tool and Domains for scoring methodological quality

Table 9. Study and participant characteristics for each included study.

Table 10. Pooled mean and standard deviation for all TROND axonal excitability indices in ALS patients and healthy controls.

Table 11. Graphical representation of methodological quality analysis.

Figure 1. Forest plot of CMAP amplitudes.

Figure 2. Forest plot of rheobase.

Figure 3. Forest plot of subexcitability.

Figure 4. Forest plot of relative refractory period (RRP).

Figure 5. Forest plot of refractoriness at 2 ms.

Figure 6. Forest plot of TEd 40-60ms

Figure 7. Forest plot of TEh 10-20ms

Figure 8. Forest plot of TEh 90-100ms

Figure 9. Forest plot of 50% depolarizing I/V index.

Figure 10. Forest plot of 100% hyperpolarizing I/V index.

Figure 11. Forest plot of resting I/V slope.

Figure 12. Forest plot of hyperpolarizing I/V slope.

Figure 13. Risk of bias and applicability concerns

## ACKNOWLEDGMENTS

We thank Dr. Meghan Sebastianski (Alberta SPOR Support Unit) for providing advice on the meta-analysis procedure. We thank Dr. Siyu Du for assistance in preparation of the revised manuscript and the graphical abstract. We would also like to thank the members of the ALS community for their input in shaping this review.

## AUTHOR CONTRIBUTIONS

KJ and AL had full access to the data set of this study and take responsibility for the integrity of data and the accuracy of data analysis. Conception and design: KJ, AL, and MS. Systematic search: AS. Articles screening: KJ, AL, MS, and HT. Data extraction, statistical analysis, and interpretation of data: KJ and AL. First draft of manuscript: AL. Critical revision of manuscript: KJ, AS, MS, and HT. Supervision: KJ. All authors made substantial contributions to the intellectual content of the paper and gave final approval for the final version of the manuscript.

